# Continuous Electroencephalography (cEEG) Characteristics and Acute Symptomatic Seizures in COVID-19 Patients

**DOI:** 10.1101/2020.05.26.20114033

**Authors:** Shreya Louis, Andrew Dhawan, Christopher Newey, Dileep Nair, Lara Jehi, Stephen Hantus, Vineet Punia

## Abstract

**Background:** Neurological manifestations of COVID-19 have only recently been described, with a paucity of literature reporting the potential relationship between COVID-19 and acute symptomatic seizures. Two prior studies found no clinical or electrographic seizures in their cohorts of COVID-19 patients with altered mental status (AMS) and clinical seizure-like events (SLEs).

**Methods:** In this retrospective cohort study, 22 critically-ill COVID-19 patients above the age of 18 years who underwent EEG (electroencephalography) monitoring between April 20th, 2020 and May 20th, 2020 were studied. 19 patients underwent continuous EEG (cEEG) for at least 24 hours, and 3 patients underwent routine EEGs (<1 hour). Demographics including age, gender, comorbid medical, and neurological conditions were collected. Clinical variables included EEG findings, anti-seizure medications, discharge disposition, and survival.

**Findings:** 17 patients underwent EEG monitoring for unexplained altered mental status changes and 5 patients underwent monitoring for a seizure-like event. 5 patients had epileptiform abnormalities on EEG (4 patients on cEEG, 1 on routine EEG); and only 2 of 5 epileptic EEG patients had a prior history of epilepsy. 2 patients in our cohort had electrographic seizures in the absence of prior epilepsy history. No patients with epileptiform abnormalities or electrographic seizures had acutely abnormal neuroimaging on CT or MRI.

**Interpretation:** Encephalopathic COVID-19 positive patients had a range of EEG abnormalities, and a higher proportion of patients in this series had electrographic seizures than previous literature suggests. This may be influenced by the duration of monitoring with cEEG and the use of a 21 channel electrode system. cEEG findings may help to guide antiseizure medical therapy, as well as the workup of altered mental status in the setting of unremarkable neuroimaging.

**Funding:** No funding was used for this study.

## Introduction

As the COVID-19 pandemic continues, there is a need to better understand the neurological manifestations of the infection and its associated complications. In association with their critical illness, patients with COVID-19 are often evaluated for seizures, but there is a limited understanding of whether seizures are a manifestation or a byproduct of the disease process itself. Acute, clinical and non-clinical seizures are well-known complications in critically ill patients with sepsis and brain injury^1-3^. Indeed, the use of continuous EEG (cEEG) monitoring in acutely ill patients, has revealed that a majority of acute seizures are exclusively non-convulsive^4,5^.

An initial retrospective case series of 214 patients by Mao et al. described CNS manifestations in 53 patients, of whom 6 patients had cerebrovascular disease and one patient had a clinical seizure^6^. A study of 304 patients by Lu et al. showed no evidence of acute symptomatic seizures and only two patients with seizure-like symptoms in the settings of hypocalcemia and acute stress^7^. One patient, as reported by Vollono et al., was noted to have status epilepticus after being otherwise seizure-free for two years on valproic acid^8^.

EEG evaluations have also been studied in COVID-19 infected patients. Galanopolou et al. described EEG findings in 22 COVID-19 positive patients. Of these, 9 patients had sharp waves, including 8 in the frontal region, and no electrographic seizures were recorded^9^. This high percentage of epileptiform abnormalities (EAs) in a specific brain region in COVID-19 patients behooves the question if this is a potential epileptic effect from the SARS-CoV-2 infection or if this is a limitation with the EEG requisition method itself, since patients did not receive the minimum required 21-electrode clinical EEG recommended by American Clinical Neurophysiological Society ACNS)^10^.

To add to the understanding of EEG findings in COVID-19 positive patients, we present a cohort of 22 COVID-19 positive patients, of whom 19 underwent continuous EEG (cEEG) with 21-electrodes for at least 24 hours, and 3 underwent routine EEG (<1 hour). All patients were recorded with a minimum of 21-electrodes. This is the largest cEEG study to date in COVID-19 patients. We determine the prevalence of EEG changes in this population, the characteristics of these EEG changes, and the relationship of EEG findings to survival.

## Methods

### Study Population

After IRB approval, we cross-matched the Cleveland Clinic COVID-19 registry with our prospectively maintained EEG database (Ebase, Cleveland, OH) from April 20th, 2020 until May 20th, 2020. All hospitalized COVID-19 positive adults (≥18 years of age at the time of diagnosis of COVID-19) who underwent an EEG were included in the study population. Patients were excluded if they had negative COVID-19 testing or if they did not undergo EEG evaluations during the time they were admitted for COVID-19 infection. We identified 19 COVID-19 positive patients who underwent cEEG for at least 24 hours and 3 COVID-19 positive patients who underwent routine EEG (20 minutes). All variables in this study were collected until May 20th 2020.

### Data Collection

COVID-19 disease status was assayed by nasopharyngeal swabs utilizing the Centers for Disease Control and Prevention (CDC) reverse transcription polymerase chain reaction (RT-PCR) SARS-CoV-2 assay (Roche Magnapure extraction and ABI 7500 DX) as validated in the Cleveland Clinic Robert J. Tomsich Pathology and Laboratory Medicine Institute.

All COVID-19 patients had at least one SARS-CoV-2 positive test prior to the initiation or during cEEG monitoring. Electronic medical record (EMR; EPIC, Verona, WI) was reviewed to extract predefined COVID-19-related clinical variables: fever, pneumonia, mechanical ventilation status, and treatment (e.g. hydroxychloroquine). Medical and neurological comorbidities, including epilepsy history and the use of anti-seizure medications (ASMs), were also extracted from EMR review. Medical comorbidities included, but were not limited to: chronic obstructive pulmonary disease (COPD), congestive heart failure (CHF), diabetes, hypertension, coronary artery disease, cancer, and immunosuppressive disease.

### Continuous EEG Monitoring

cEEG at our institution uses 21 electrodes placed according to the international 10-20 system by certified EEG technologists. Every study patient underwent monitoring for at least 24 hours and cEEG tracings were interpreted by board-certified neurophysiologists with extensive experience in interpreting cEEGs. EEG findings were extracted from the EEG reporting database. EEG findings were classified using the ACNS terminology for cEEG^11^ Electrographic seizures were classified based on Salzburg criteria^12^. Epileptiform abnormalities (EAs) included isolated epileptiform discharges e.g. sharp waves/ spikes/polyspikes^13^, lateralized periodic discharges (LPDs, formerly PLEDs)^14^, and lateralized rhythmic delta activity (LRDA)^15^. We used ILAE (International League Against Epilepsy) terminology for description of ictal semiology associated with electrographic seizures^16^.

cEEG monitoring indications were collected from the history portion of the EEG requisition form. EMR was reviewed in cases where the above did not clarify the indication for cEEG monitoring. Indications were coded as either unexplained altered mental status (AMS) or seizure-like event (SLE). Unexplained AMS was defined by the treating clinician in all cases, representing a change from baseline mentation not accounted for by the patient’s medical condition or drugs, leading to a clinical concern for non-convulsive seizures (NCS). SLEs were also defined by the treating clinician and primarily represented motor events such as clonic or myoclonic movements. Patients with AMS after a witnessed SLE were classified into the latter category.

### Clinical Outcomes Data Collection

While the main goal of this study was to retrospectively report cEEG findings in a cohort of critically ill COVID-19 positive patients, we also explored putative associations between survival, clinical outcomes (discharge disposition), cohort characteristics (e.g. comorbidities, age, gender), and EEG findings.

### Statistical Analysis

Continuous and categorical data were summarized with mean values, standard deviations, medians (continuous data), and frequencies (categorical data). The Shapiro Wilks test assessed normality, and based on the distribution of continuous variables, the Wilcoxon rank sum test or student’s t-test was performed. Fisher’s exact test was used for categorical variables to compare cohort characteristics between patients who expired in the hospital with patients who are still alive. All analysis was performed using R statistical software v3.6.3 (Foundation for Statistical Computing, Vienna, Austria)^17^ and all tables were created using the Table1 R package v1.2 (Rich 2020)^18^. Bonferroni correction was used for multiple test correction in Table 2, no multiple test correction was performed on the demographics shown in Table 1. A p value of 0.05 or less was considered significant in the demographics table, and the Bonferroni adjusted cutoff p value was 0.005 for Table 2.

**Table 1:** Cohort Characteristics.

|  | **Total**<br>**(N=22)** | **Alive**<br>**(N=16)** | **Expired in Hospital**<br>**(N=6)** | **p** |
| --- | --- | --- | --- | --- |
| **Sex** |  |  |  | > 0.999 |
| *Female* | 8 (36.4%) | 6 (37.5%) | 2 (33.3%) |  |
| *Male* | 14 (63.6%) | 10 (62.5%) | 4 (66.7%) |  |
| **Age (years)** |  |  |  | **0.023\*** |
| *Mean (SD)* | 66.5 (11.2) | 63.4 (10.7) | 74.8 (8.40) |  |
| **Race** |  |  |  | 0.360 |
| *African American* | 7 (31.8%) | 5 (31.2%) | 2 (33.3%) |  |
| *Asian* | 1 (4.5%) | 0 (0%) | 1 (16.7%) |  |
| *White* | 14 (63.6%) | 11 (68.8%) | 3 (50.0%) |  |
| **History of non-neurological comorbidities** |  |  |  |  |
| *Smoking* | 10 (45.5%) | 7 (43.8%) | 3 (50.0%) | > 0.999 |
| *Chronic obstructive pulmonary disease* | 2 (9.1%) | 1 (6.2%) | 1 (16.7%) | 0.481 |
| *Asthma* | 8 (36.4%) | 6 (37.5%) | 2 (33.3%) | > 0.999 |
| *Diabetes* | 9 (40.9%) | 6 (37.5%) | 3 (50.0%) | 0.655 |
| *Hypertension* | 15 (68.2%) | 10 (62.5%) | 5 (83.3%) | 0.616 |
| *Coronary artery disease* | 4 (18.2%) | 1 (6.2%) | 3 (50.0%) | **0.046\*** |
| *Congestive heart failure* | 4 (18.2%) | 2 (12.5%) | 2 (33.3%) | 0.668 |
| *Cancer* | 7 (31.8%) | 4 (25.0%) | 3 (50.0%) | 0.530 |
| *Immunosuppressive disease* | 8 (36.4%) | 4 (25.0%) | 4 (66.7%) | 0.137 |
| **History of neurological comorbidities** | 6 (27.3%) | 4 (25.0%) | 2 (33.3%) | > 0.999 |
| *Epilepsy* | 2 (9.1%) | 2 (12.5%) | 0 (0%) | > 0.999 |
| *Stroke* | 1 (4.5%) | 0 (0%) | 1 (16.7%) | 0.273 |
| *Headache* | 1 (4.5%) | 0 (0%) | 1 (16.7%) | 0.273 |
| *Traumatic brain injury* | 1 (4.5%) | 1 (6.2%) | 0 (0%) | > 0.999 |
| *Spinal stenosis* | 2 (9.1%) | 2 (12.5%) | 0 (0%) | > 0.999 |
| **COVID-19 specific characteristics** |  |  |  |  |
| *Fever* | 15 (68.2%) | 12 (75.0%) | 3 (50.0%) | 0.334 |
| *Pneumonia* | 6 (27.3%) | 4 (25.0%) | 2 (33.3%) | > 0.999 |
| *Mechanical ventilation required* | 18 (81.8%) | 12 (75.0%) | 6 (100%) | 0.541 |
| *Hydroxychloroquine given as treatment* | 16 (72.7%) | 11 (68.8%) | 5 (83.3%) | 0.634 |

p values reported from Wilcoxon Rank Sum test for continuous variables and Fisher's exact for categorical variables.
p value 0.01-0.05 - \*, p value 0.001 to 0.05 - \*\*

**Table 2:**
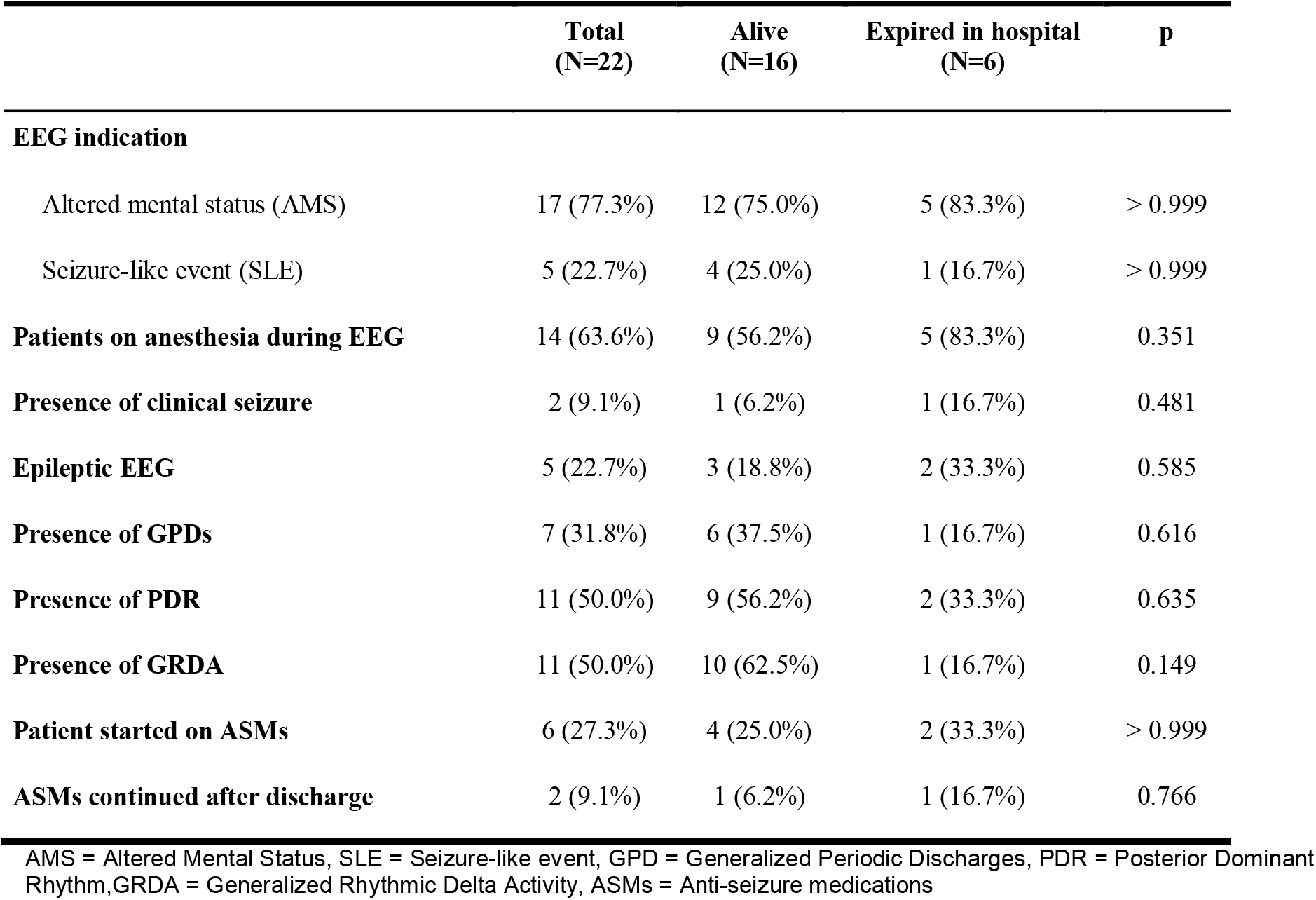
Cohort EEG Findings by Outcome.

## Results

### Cohort demographics and comorbidities

A total of 22 COVID-19 patients (8 females; 36.4%), with a mean age of 66.5 years (±11.2 years), underwent EEG monitoring. All except 3 patients underwent cEEG monitoring. Demographics, medical and neurological comorbidities, as well as COVID-19 specific characteristics of the study population are summarized in **Table 1**. Briefly, 15 patients (68.2%) presented with fever at some point in their hospital admission, 6 patients (27.3%) developed pneumonia, and 18 patients (81.8%) required mechanical ventilation (**Table 1**). Of the full cohort, 16 patients (72.7%) received hydroxychloroquine during their hospitalization. Neurological comorbidities were present in 27.3% of patients. Only one patient had a prior history of stroke (4.8%), and two (9.1%) patients had a known history of epilepsy (**Table 1**).

### Clinical Outcomes

At the point of most recent follow-up (May 20th 2020), 6 patients (27.3%) did not survive hospitalization, and the remainder were discharged (16 patients). The patients who expired were significantly older (mean 74.8 ± 8.4 years) than those who survived during the course of hospitalization (mean 63.4 ± 10.7 years; p = 0.023). The length of hospital stay ranged from 3 to 36 days. There was no statistically significant difference in the hospital length of stay between those who expired in hospital (mean 16.7 ± 4.6 days) and those who survived to last follow-up (mean 20.8 ± 11.1 days; p = 0.226). Of the 16 patients who were discharged, 4 were discharged to their respective homes, 6 were sent to skilled nursing facilities (SNF), 3 were sent to long term care facilities, 2 were sent to inpatient rehabilitation facilities and 1 was sent to an acute care hospital.

### cEEG Findings and Outcomes

Among the study population, 19 (86.4%) patients underwent cEEG and 3 patients had routine EEGs. The summary of EEG and clinical variables of the study cohort are provided in Table 2. Of 19 cEEG patients, 12 (63.2%) were on IV infusions of anesthetics (fentanyl, propofol, and/or midazolam) during at least a portion of cEEG monitoring. The median duration of cEEG monitoring was 2 days (range 1 - 6 days). In 4 of 19 patients, cEEG was performed for evaluation of seizure-like events (SLE). These were described as left arm clonic movements, right face twitching (clonic), witnessed generalized tonic clonic seizure, and “eyebrow twitching with lip smacking.” The remaining 15 patients underwent cEEG evaluation to rule out non-convulsive seizures (NCS) as a cause of unexplained altered mental status AMS.

Acute electrographic seizures were noted in 2 (10.5%) of the 19 cEEG patients. Epileptic abnormalities were noted in 4 of 19 cEEG patients (15.7%), including the 2 with electrographic seizures. The remaining 2 patients had a history of epilepsy and were noted to have generalized polyspikes and right frontal sharp waves.

The cEEG showed a continuous (>80% of the recording) generalized polymorphic delta slowing in all patients. The posterior dominant rhythm (PDR) was absent throughout the cEEG in 8 patients (42.1%), was slow (<8 Hz) in 9 patients (47.4%), and within normal limits in the remaining patients. Six of 8 patients lacking PDR on cEEG were on IV anesthetics at some point during the cEEG monitoring. The proportion of patients on IV anesthetics was not statistically different in the patients lacking PDR compared to ones with a discernible PDR (54.5%, p=0.633). Generalized periodic discharges (GPDs) were noted in 7 (36.8%) patients, ranging from 0.5 Hz to 1 Hz in frequency. In 5 of these patients, GPDs were of triphasic morphology and in two, the GPDs were sharply contoured. Ten patients (52.6%) were noted to have intermittent generalized rhythmic delta activity (GRDA), including 3 patients with sharply contoured waveforms.

Six patients (27.3%) were started on an anti-seizure medication (ASM); levetiracetam was used in five patients, and valproic acid in one patient. Of these six patients, only three patients were found to have epileptic EEGs. At discharge, there was one patient who continued on levetiracetam 500 mg BID due to the presence of epileptic EEG findings. Another patient who was started on Levetiracetam due to abnormal EEG findings expired in hospital (Case 2 below). A third patient had no presence of epileptic EEG findings, but was discharged on Valproate 500 mg.

### Routine EEG Findings

There were 3 patients (one female) of the 22 who underwent routine EEGs. None had a prior history of epilepsy. Of these three patients, two were receiving IV anesthesia at the time of routine EEG monitoring. Two patients were monitored for unexplained AMS, and one patient underwent EEG due to a SLE of “right-sided jerking movements.” The patient with the SLE had an epileptic EEG showing left hemispheric LRDA up to 1.5 Hz in frequency. None of the three patients with routine EEG had PDRs.

With respect to clinical outcomes, two of the three patients expired in the hospital, and one was discharged home.

### Description of acute symptomatic seizures

There were 2 patients with acute symptomatic clinical seizures that have been reported elsewhere. They were both on cEEG. None of them had an epilepsy history.

Case 1:

Patient was a 76 year old white male with a history of asthma, hypertension, diastolic heart failure, non-epithelial skin cancer, and lumbar stenosis. He was admitted for encephalopathy and a fever (39.2°C), five days after a lumbar spine surgery with concern for an epidural abscess, drained on the day of admission. Abscess cultures grew Pseudomonas, and vancomycin with piperacillin-tazobactam was started. He developed acute hypoxemic respiratory failure with high fever, and was transferred to the intensive care unit. On postoperative day two, the patient was noted to have several episodes of left upper extremity clonic movement with worsening encephalopathy. cEEG was started and captured three left arm clonic seizures lasting for roughly 30 seconds each, originating from the right centroparietal region. Levetiracetam was initiated with subsequent cessation of clinical and electrographic seizures. MRI brain with and without contrast only noted chronic white matter hyperintensities, and no focal lesion. Due to persistently high fever, acyclovir was added to his antimicrobial regimen, and a respiratory viral panel was sent, along with COVID-19 testing. COVID-19 testing returned positive, and acyclovir was discontinued. Ultimately, after 30-days of hospitalization, the patient was discharged to SNF on levetiracetam.

Case 2:

Patient was an 82 year old African-American male with a past medical history significant for smoking, COPD, asthma, hypertension, coronary artery disease, heart failure, and prostate cancer. He was admitted for progressive dyspnea over the preceding ten days, generalized weakness, and altered mental status. He was hypoxic, febrile and tachycardic at time of admission, requiring intubation and mechanical ventilation. He tested positive for COVID-19. cEEG was initiated due to the presence of right eyelid and facial twitching on admission day 5. Multiregional EEG seizures arising from the left and right fronto- temporal regions (left greater than right) and left parieto-occipital region were captured. Most seizures were non-convulsive, but 4 episodes of clonic facial movements associated with fronto-temporal seizures were also noted. Seizure frequency significantly improved after treatment with levetiracetam. A non-contrast CT brain did not reveal any acute intracranial findings. Lumbar puncture could not be performed due to coagulopathy. Unfortunately, this patient expired in the hospital 17 days into his admission.

### EEG Findings Do Not Show Statistically Significant Association with Survival

Among the 22 patients in the study population there were no statistically significant differences between patients who expired in the hospital and those who remain alive with respect to the presence of epileptic EEG findings (p=0.585), PDR (p=0.635), GPD (0.616), or GRDA (p = 0.149) (Table 2).

### Neuroimaging Did Not Have EEG Correlate

Most of the study population (18 patients, 85.7%) received neuroimaging in the form of a non-contrast CT brain. The majority of CT brain imaging revealed no acute intracranial processes. One patient had an intracerebral hemorrhage; one patient had an acute ischemic stroke, and two patients had imaging concerning for possible ischemia. MRI brain was obtained in five patients (23.8%). Of these, one had an MRI within normal limits, one showed an acute infarct (as suspected on CT brain), one showed chronic white matter hyperintensity (Case 1, above), another had a remote hemorrhage, and one had a remote infarct. Except for Case 1 with acute symptomatic seizure, no patients with the listed CT/MRI findings had EAs or electrographic seizures recorded on EEG.

### Discussion

In this work, we present a comprehensive description of EEG findings in COVID-19 infected patients. Excluding the 2 patients with history of epilepsy, 3 of 20 patients had epileptic findings. Among them, one patient was found to have left LRDA (as epileptic as LPDs in critically ill patients, Gaspard et al.)^23^, and two others had electrographic seizures. All cEEGs showed changes consistent with encephalopathy in the form of continuous slowing in the delta frequency range. While the contribution of IV anesthetics to cEEG findings may confound the analysis, no statistically significant difference in rate of PDR was seen between those patients on or off of IV anesthetics. Slightly more than half of our cEEG patients were found to have GRDA, a finding known to be non-epileptic, and another one-third (36.8%) were found to have GPDs. While GPDs may be epileptic and associated with electrographic seizures, the relationship is frequency dependent^5,19^. Two patients had sharply contoured GPDs, both with afrequency less than 1 Hz, below the 1.5 Hz threshold known to correlate with increased seizure risk^5^. Galanopoulou et al. found GPDs in only 1 out of 22 (4.5%) COVID-19 patients 14, lower than what was observed in our study. Notably, this difference may have arisen due to a lower rate of cEEG usage in their cohort (7 of 22 patients).

Sporadic inter-ictal epileptiform discharges (IEDs) such as sharp waves were not seen in patients lacking a history of epilepsy. This lack of sharp waves is in contrast to 9 out of 22 patients found to have sharp waves in the work by Galanopoulou et al.^14^, in which all but one had frontal sharp waves. Although the duration of 8 channel-EEGs performed in the study is unclear, these are typically used for screening purposes and not for continuous use. In contrast, cEEG patients in this study underwent a median duration of 2 days of monitoring. Although the severity of illness in the two patient populations cannot be compared, the most likely explanation of this major difference in the EEG findings is the lack of a 10-20 EEG system using the minimum required 21 electrodes^10^. As acknowledged by authors and the American Clinical Neurophysiological Society, the chances of interpretive errors increase with fewer electrodes; this is particularly the case for transient findings such as sharp waves.

So far, clearly documented epileptic seizures in COVID-19 patients, without pre-existing epilepsy, have been extremely rare. Indeed, an individual found to have COVID-19 RNA in CSF with a meningo- encephalitic presentation was clinically noted to have a one-minute “generalized seizure” and “multiple epiletic seizures” requiring intubation^20^. Acute symptomatic seizures were not observed in a dedicated multicenter series to assess seizure risk in more than 300 COVID-19 individuals, nor were they evident in the case series by Galanopoulou et al. from New York^7,9^. As such, the two COVID-19 patients with clinical acute symptomatic seizures captured on EEG from our cohort are rare findings.

Both patients with acute symptomatic seizures in this study were elderly males with several comorbidities. In these patients, the lack of acute neuroimaging findings in both raises the suspicion of whether there is a direct or indirect contribution of COVID-19 infection in causing acute seizures. Whether the seizures are evidence of its neurotropism, a symptom of microthrombic events in the brain, secondary to hypoxic and inflammatory processes, or multifactorial remains unclear^21-24^. Further, while most seizures in critically ill patients are non-convulsive,^4,5^ it is curious that we report convulsive, focal motor seizures in 2 patients, perhaps also corroborated with the case reported by Vollono et al. of a patient with well-controlled epilepsy of over two years whose first manifestation of COVID-19 was myoclonic status epilepticus^8^.

In our cohort, just over a quarter (27.3%) of patients were started on ASMs, and only 2 were eventually discharged on them. In these patients, cEEG helped to clarify the lack of indication for continuation of ASM. In studies without strong use of cEEG monitoring, there is a higher rate of ASM usage, as in Galanopoulou et al., where more than 50% of patients were started on ASM.

There are several limitations of our study, which include its retrospective design and small study population. Since the primary aim of our study was characterization of cEEG findings in patients with COVID-19, we did not include patients with clinical concerns for COVID-19 disease who tested negative or matched controls. Ultimately, multicenter collaborative efforts are needed to better characterize the risk of acute and long term epileptogenic potential following COVID-19 infection, and understanding of the neurotropic capabilities of this pathogen are needed.

In conclusion, COVID-19 positive patients who were encephalopathic had a variety of epileptiform abnormalities on EEG, and a higher proportion of patients had electrographic seizures than reported in previous studies. In sharing our experience, it is our hope that cEEG monitoring can be utilized as a resource for medical decision making for ASMs, and also to better understand this disease.

## Data Availability

The data used in this study is not publically available.

